# Serum levels of specialised pro-resolving molecule pathways are greatly increased in SARS-CoV-2 patients and correlate with markers of the adaptive immune response

**DOI:** 10.1101/2021.12.07.21267409

**Authors:** James Turnbull, Rakesh Jha, Catherine A. Ortori, Eleanor Lunt, Patrick J. Tighe, William L. Irving, Sameer A. Gohir, Dong-Hyun Kim, Ana M. Valdes, Alexander W. Tarr, David A. Barrett, Victoria Chapman

**Author notes:** **Correspondence:** Professor Victoria Chapman, School of Life Sciences, University of Nottingham, University Park, Nottingham, NG7 2RD. Joint First Author (James Turnbull, Rakesh Jha).

## Abstract

**Background:** Specialised pro-resolution molecules (SPMs) halt the transition to chronic pathogenic inflammation. We aimed to quantify serum levels of pro- and anti-inflammatory bioactive lipids in SARS-CoV-2 patients, and to identify potential relationships with innate responses and clinical outcome.

**Methods:** Serum from 50 hospital admitted inpatients (22 female, 28 male) with confirmed symptomatic SARS-CoV-2 infection and 94 age and sex matched cohort collected prior to the pandemic, were processed for quantification of bioactive lipids. Anti-nucleocapsid and anti-spike quantitative binding assays were performed.

**Results:** SARS-CoV-2 serum had significantly higher concentrations of omega-6 derived pro-inflammatory lipids and omega-6 and omega-3 derived SPMs, compared to age and sex matched controls. Levels of SPMs were not markedly altered by age. There were significant positive correlations between SPMs and other bioactive lipids and anti-spike antibody binding. Levels of some SPMs were significantly higher in patients with an anti-spike antibody value >0.5. Levels of linoleic acid (LA) and 5,6-dihydroxy-8Z,11Z,14Z-eicosatrienoic acid (5,6-DHET) were significantly lower in SARS-COV-2 patients who died.

**Discussion:** SARS-COV-2 infection was associated with a robust activation of the pathways that generate the specialised pro-resolution molecules and other anti-inflammatory bioactive lipids, supporting the future investigation of these pathways which may inform the development of novel treatments.

## Introduction

Infection with severe acute respiratory syndrome coronavirus 2 (SARS-CoV-2) is characterised by fever and cough, with more severe cases developing acute respiratory distress, acute lung injury, pneumonia, and mortality[1-3]. The higher rates of severe SARS-CoV-2 illness and death are associated with increasing age[4, 5]. SARS-CoV-2 infection is associated with changes in adaptive and innate immunity, including elevated levels of circulating neutrophils and the presence of peripherally derived macrophages in the lungs of severe cases[6, 7], reduced numbers of circulating T cells[8] and robust cytokine responses, which continues after clearance of the virus[9]. SARS-CoV-2 infection is associated with elevated levels of pro-inflammatory cytokines, including IL6, IL1β, and TNFα[10, 11], however levels of anti-inflammatory cytokines IL4 and IL10 are also elevated[10]. Knowledge of the impact of SARS-CoV-2 infection on the resolution of inflammation pathways will provide crucial new mechanistic insight and potential new avenues for treatment[12-14].

Prostaglandins (PGs) and leukotrienes have essential roles in initiating acute inflammatory responses and the generation of pro-inflammatory cytokines, which sustain chronic inflammatory responses. In concert with these cyclooxygenase (COX) pathways, the lipoxygenase (LOX) pathways produce pro-inflammatory hydroxyeicosatetraenoic acids (HETEs) from arachidonic acid (AA) and hydroxyoctadecadienoic acids (HODEs) from di-homo-γ-linolenic acid (LNA). The active curtailing of inflammatory signalling is essential to restore tissue homeostasis and prevent chronic inflammatory events leading to pathology[15]. Following the initial acute inflammatory phase, specialised pro-resolving molecules (SPMs), derived from key polyunsaturated fatty acids (PUFAs), are generated and orchestrate the resolution of inflammation by promoting macrophage mediated clearance of cellular debris and counteracting the effects of pro-inflammatory cytokines[15]. The SPMs are derived from omega-6 (ω-6) (LNA, AA) or omega-3 (ω-3) (eicosapentaenoic acid (EPA), docosahexaenoic acid (DHA)) substrates via the COX, LOX and cytochrome P450 pathways[15] (SI Figure 1).

**Figure 1.**
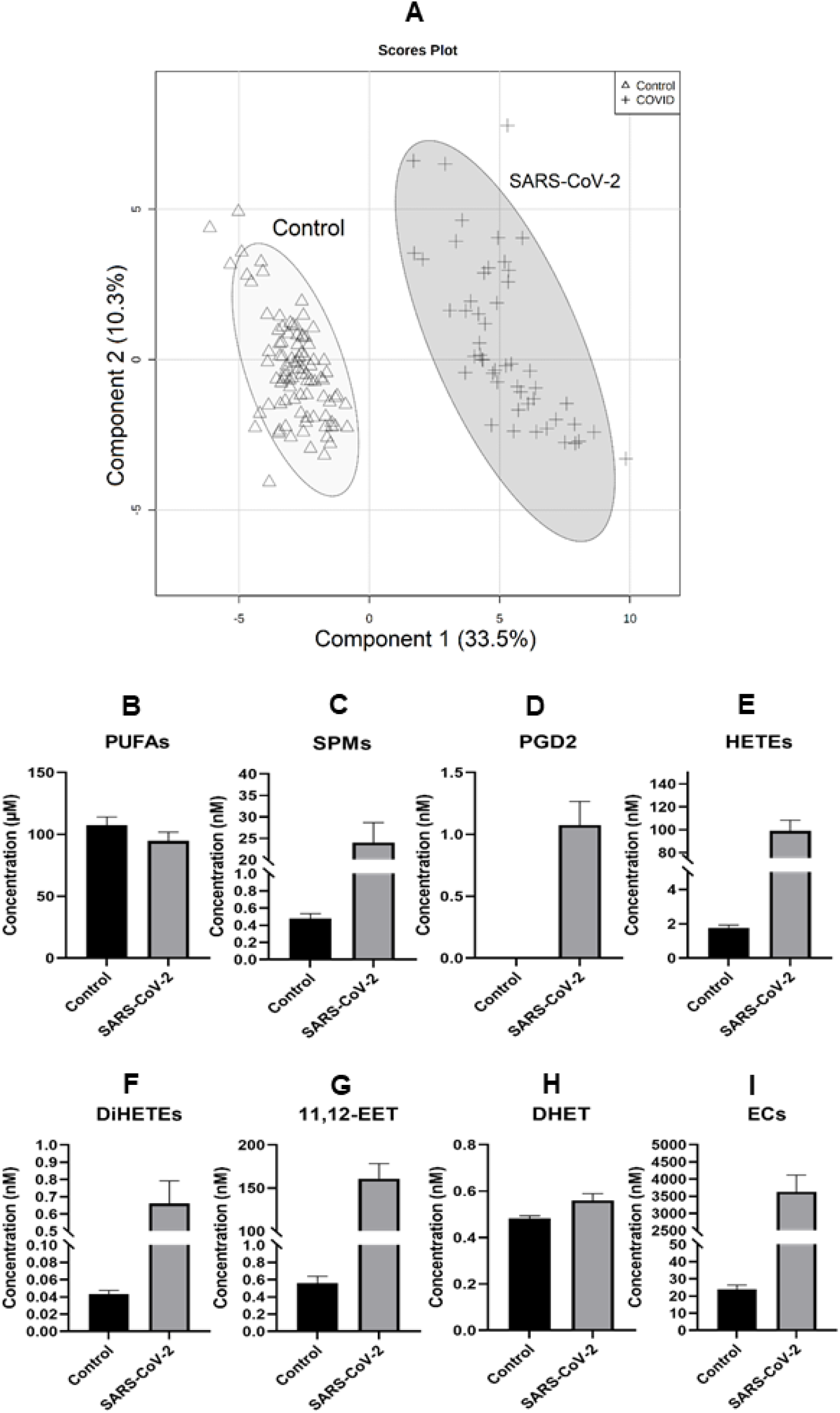
A: Partial least square discrimination analysis (PLS-DA) for the 44 serum lipids quantified in SARS-CoV-2(n=50) and age and sex matched controls (n=94) with R2 = 0.944, Q2 = 0.932 and Accuracy = 1.0 B-I: Histograms of the highest ranked lipid mediators, PUFA (AA, LA, EPA, DHA), SPMs (17-HDHA, RvD4, LXA4, LXA5), PGD2, HETEs (5-HETE, 9-HETE, 16-HETE, 19-HETE, 15-HETE), DiHETEs (5, 15-DiHETEs, 8, 9-DiHETEs), 11,12-EET, DHETs (11, 12-DHET, 14, 15-DHET) and Endocannabinoids (2-AG, AEA).

**Figure 2.**
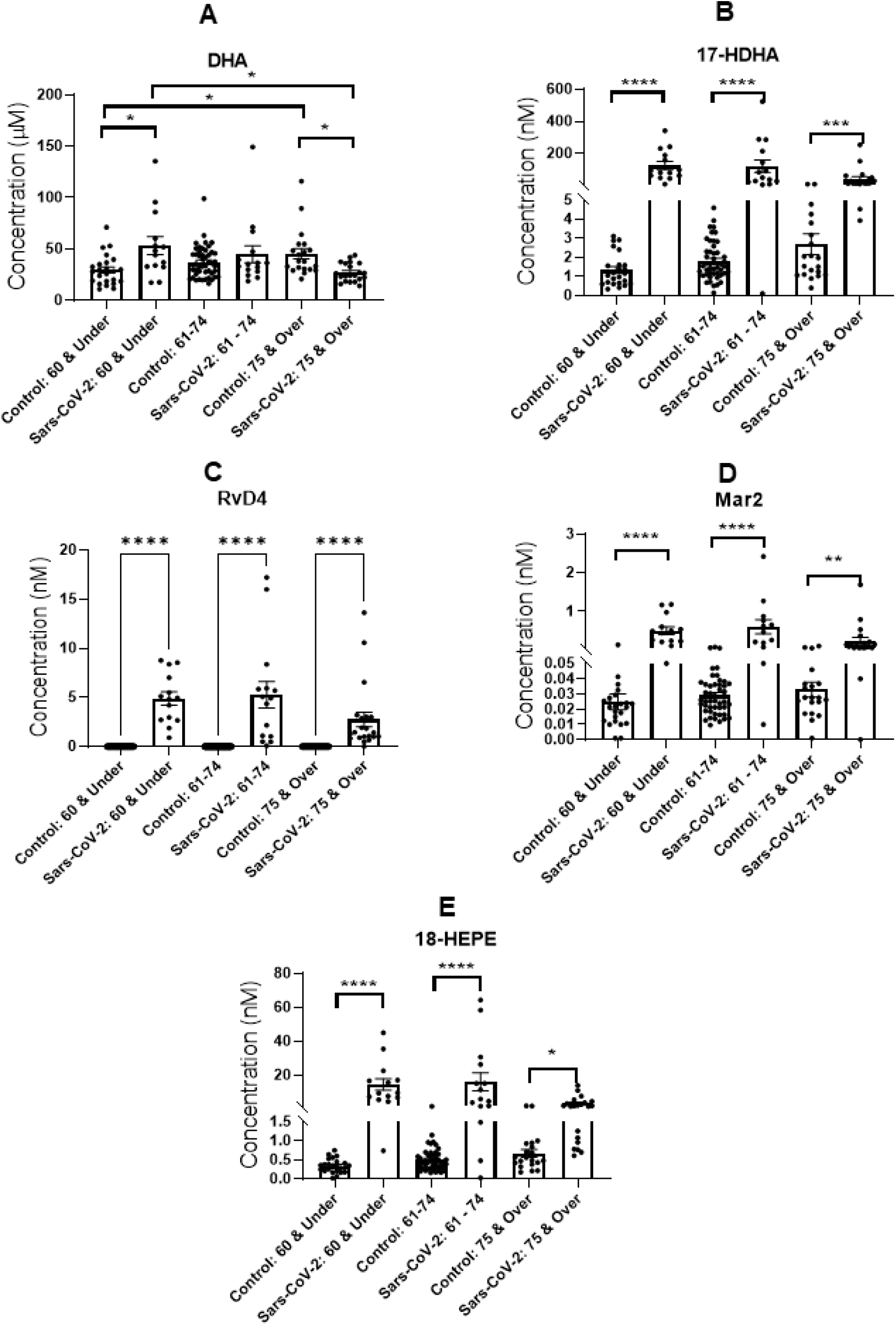
Serum concentrations of (A) DHA, (B) 17-HDHA, (C) RvD4 (D) Maresin 2, and (E) 18-HEPE in SARS-CoV-2 (n=50) and age and sex matched control serum (n=94) stratified by age group. Groups were assessed for normal distribution using D’Agostino & Pearson test. Significance was assessed using Kruskal-Wallis test correcting for multiple comparisons using Dunn’s test. *p=<0.05, **p=<0.01, ***p=<0.001, ****p=<0.0001.

The most well characterised SPMs are the resolvins (Rvs), protectins (PDs), and maresins (MaRs), which halt the transition from acute to chronic inflammation preventing pathogenesis[16]. 17(S)-hydroxy Docosahexaenoic acid (17-HDHA), a substrate for the generation of the D series resolvins[15], enhances the adaptive immune response in a preclinical model of influenza[17], both 17-HDHA and resolvin D1 enhance B cell production and promote B cell differentiation towards an antibody secreting phenotype[18]). The epoxyeicosatrienoic acids (EETs), derived from AA via soluble epoxide hydrolase (sEH), also mediate resolution of inflammation [19, 20], down-regulating inflammatory transcription factors such as NF-kB [21], curtailing the induction of COX2 and production of cytokines[22].

Current knowledge of the consequences of SARS-CoV-2 infection on endogenous levels of SPMs and EETs is in its infancy. Building a comprehensive picture of the impact of SARS-CoV-2 infection upon the serum lipidomic profile will aid understanding of the therapeutic potential of resolution pathways for SARS-COV-2 infection[12, 13, 23]. Our aims were to 1) compare serum levels of a range of SPMs and pro-inflammatory bioactive lipids between patients admitted to hospital with SARS-CoV-2 infection and an age-matched control group, 2) determine the potential relationship between levels of these bioactive lipids and levels of anti-nucleocapsid and anti-spike antibody titre, markers of the production of an adaptive immune response[24] and 3) investigate outcomes following infection.

## Methods

### Sample Collection and Preparation

Serum samples obtained from 50 inpatients admitted to Nottingham University Hospitals NHS Trust Queen’s Medical Centre with symptomatic RT-PCR confirmed SARS-CoV-2 infection were collected as diagnostic specimens for clinical chemistry testing, excess sera were provided anonymously for research purposes. Review by the University of Nottingham’s School of Life Sciences Ethical Review Committee deemed the study to not require full ethical review. Approval for use of anonymized clinical data was provided by the NHS Health Research Authority (HRA) and Health and Care Research Wales (HCRW) (ref. 20/HRA/4843). Samples were determined not to be Relevant Materials in line with the Human Tissue Authority. Risk assessments were approved by the UK Health and Safety Executive (ref. CBA1.470.20.1). Serum samples were initially stored at 4°C for 24 h and then inactivated with the WHO-approved protocol (4-hour room temperature incubation with 1% Triton X-100 in PBS) before analysis. Detailed methods on the assessment of the potential effect of the viral deactivation protocol on serum lipid levels are detailed in the supplementary information. Viral genomic sequencing of samples from a subset of these patients was performed as part of the COG-UK consortium[25].

Baseline serum samples from the iBEAT-OA cohort study (n=94) were used as age and sex matched controls[26], which were collected prior to the SARS-CoV-2 pandemic served as a SARS-CoV-2 negative control. As collected pre-pandemic a lack of SARS-CoV-2 infection was not confirmed. The iBEAT-OA cohort had a confirmed diagnosis of osteoarthritis. Ethical approval was obtained from the Research Ethics Committee (ref: 18/EM/0154) and the Health Research Authority (protocol no: 18021).

### Lipidomic Analysis

Serum bioactive lipids were extracted and measured using our published liquid chromatography-mass spectrometry (LC-MS/MS) quantification method for the major classes of pro- and anti-inflammatory lipid molecules, which has been updated to include SPMs and their precursor molecules[27]. 44 bioactive lipids were quantified, detailed methods are in Supplementary Information.

### Anti-Nucleocapsid & Anti-Spike Binding Assays

Anti-nucleocapsid and anti-spike quantitative binding assays were performed on serum samples using ELISA following the protocol described by Tighe et al (2020)[24]. Levels of C reactive protein (CRP) were measured in clinical diagnostic tests (mean 172.9 levels ranged from 11-489). The median time between sample collection for CRP measurement and collection of serum for the measurement of bioactive lipids, anti-nucleocapsid and anti-spike antibody binding was 5 days.

### Data Analysis

GraphPad Prism (Version 8.2.1) was used. In some cases, patients were stratified into three age groups (<60, 61-74, & >75 years). Groups were assessed for normal distribution using D’Agostino & Pearson test and evaluated for significant changes between groups using Kruskal-Wallis test with multiple comparisons corrected for using Dunn’s test. Multivariate analysis using Metaboanalyst 4.0 (https://www.metaboanalyst.ca/)[28] including principal component analysis (PCA) was performed. Partial least square discrimination analysis (PLS-DA) was used to identify the lipid mediator clusters. A cross validation analysis was used to validate the PLS-DA model based on accuracy, R2 and Q2 scores and a permutation test (SI Figure 3). Variable Importance in Projection (VIP) scores greater than 1 were recognized as playing a key role in cluster differentiation (SI Figure 4 & SI Table 1).

**Table 1.**
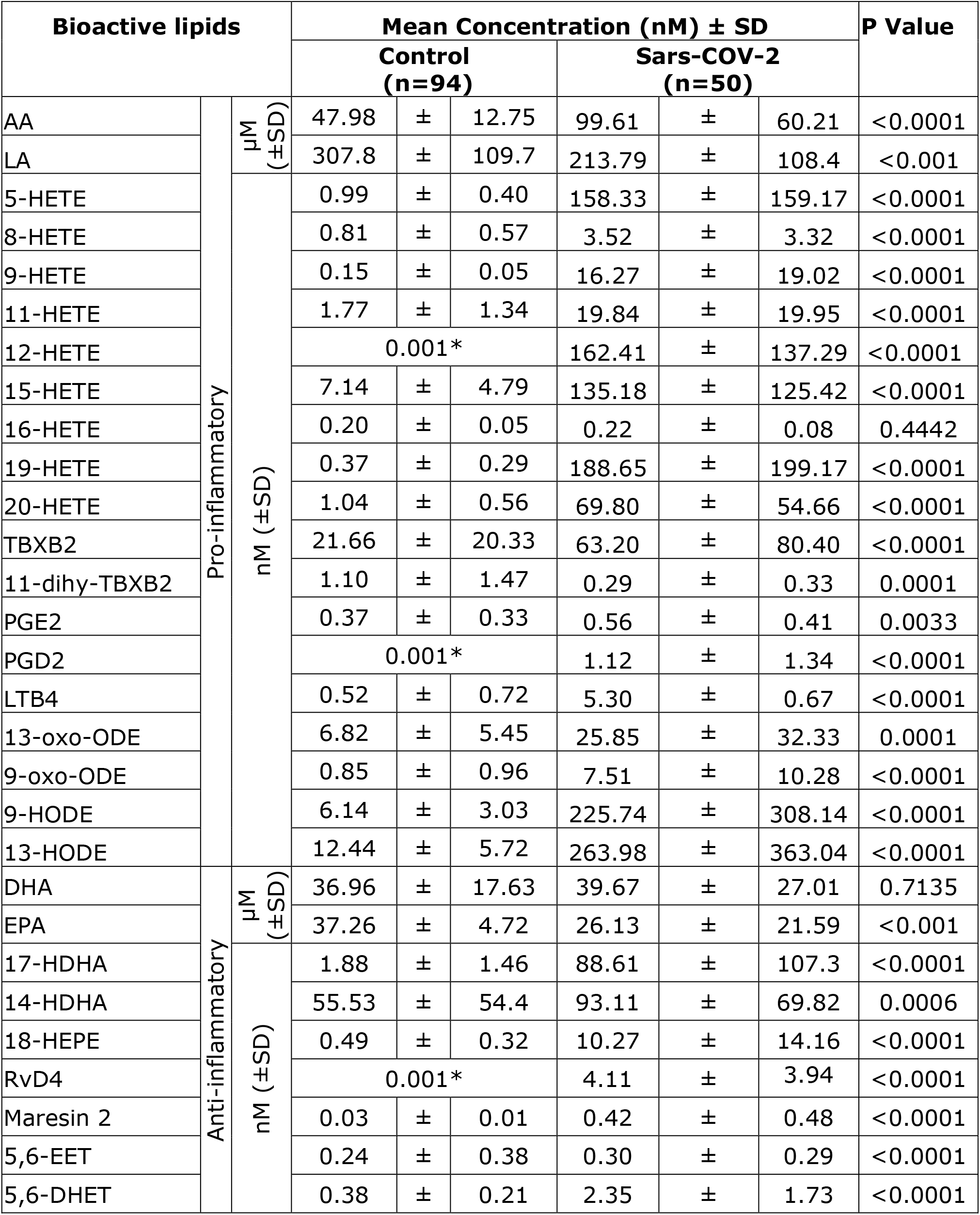
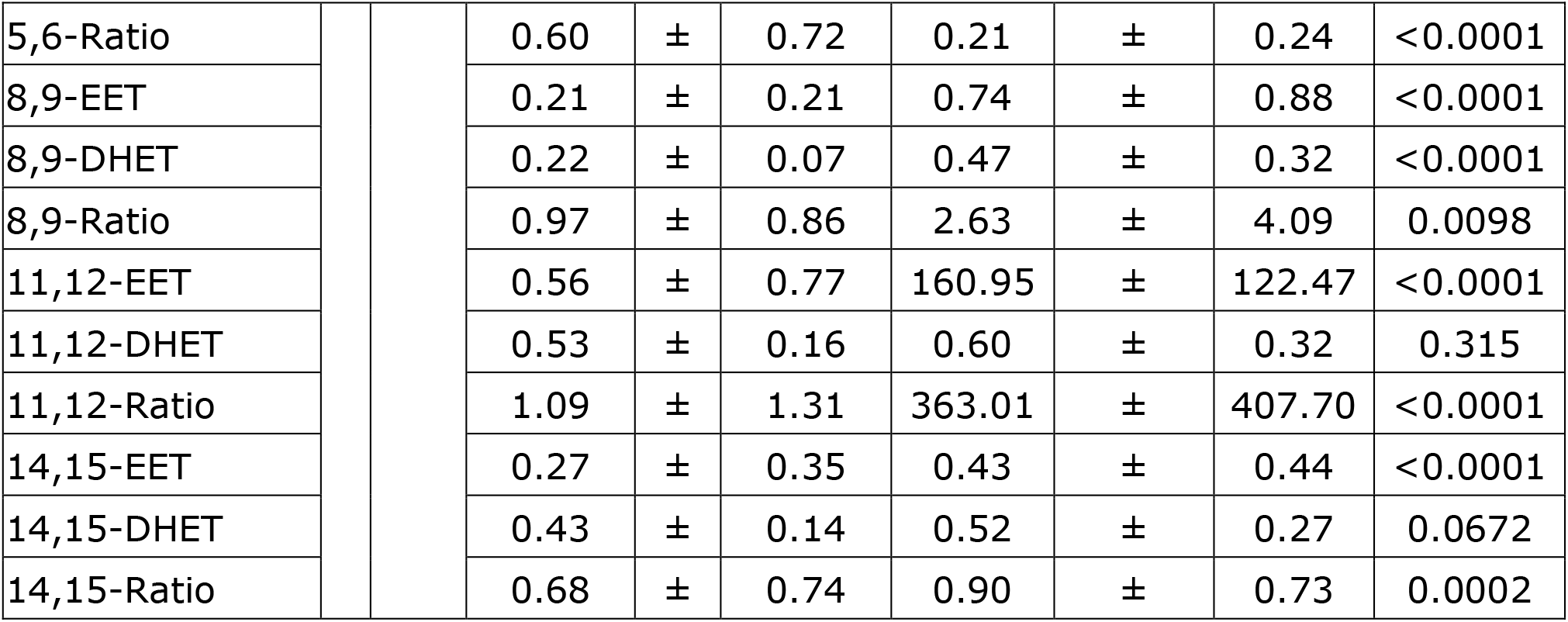
Concentrations of pro-inflammatory and anti-inflammatory bioactive lipids quantified in SARS-CoV-2 (n=50) and age and sex matched controls (n=94). Statistical analysis by Mann-Whitney Test. Where lipids were not detected in samples, an arbitrary value of 0.001 indicated by * was used for statistical analysis.

| Bioactive lipids |  |  | Mean Concentration (nM) ± SD |  |  |  |  |  | P Value |
| --- | --- | --- | --- | --- | --- | --- | --- | --- | --- |
|  |  |  | Control<br>(n=94) |  |  | Sars-COV-2<br>(n=50) |  |  |  |
| AA | Pro-inflammatory | μM<br>(±SD) | 47.98 | ± | 12.75 | 99.61 | ± | 60.21 | <0.0001 |
| LA |  |  | 307.8 | ± | 109.7 | 213.79 | ± | 108.4 | <0.001 |
| 5-HETE |  | nM (±SD) | 0.99 | ± | 0.40 | 158.33 | ± | 159.17 | <0.0001 |
| 8-HETE |  |  | 0.81 | ± | 0.57 | 3.52 | ± | 3.32 | <0.0001 |
| 9-HETE |  |  | 0.15 | ± | 0.05 | 16.27 | ± | 19.02 | <0.0001 |
| 11-HETE |  |  | 1.77 | ± | 1.34 | 19.84 | ± | 19.95 | <0.0001 |
| 12-HETE |  |  | 0.001* |  |  | 162.41 | ± | 137.29 | <0.0001 |
| 15-HETE |  |  | 7.14 | ± | 4.79 | 135.18 | ± | 125.42 | <0.0001 |
| 16-HETE |  |  | 0.20 | ± | 0.05 | 0.22 | ± | 0.08 | 0.4442 |
| 19-HETE |  |  | 0.37 | ± | 0.29 | 188.65 | ± | 199.17 | <0.0001 |
| 20-HETE |  |  | 1.04 | ± | 0.56 | 69.80 | ± | 54.66 | <0.0001 |
| TBXB2 |  |  | 21.66 | ± | 20.33 | 63.20 | ± | 80.40 | <0.0001 |
| 11-dihy-TBXB2 |  |  | 1.10 | ± | 1.47 | 0.29 | ± | 0.33 | 0.0001 |
| PGE2 |  |  | 0.37 | ± | 0.33 | 0.56 | ± | 0.41 | 0.0033 |
| PGD2 |  |  | 0.001* |  |  | 1.12 | ± | 1.34 | <0.0001 |
| LTB4 |  |  | 0.52 | ± | 0.72 | 5.30 | ± | 0.67 | <0.0001 |
| 13-oxo-ODE |  |  | 6.82 | ± | 5.45 | 25.85 | ± | 32.33 | 0.0001 |
| 9-oxo-ODE |  |  | 0.85 | ± | 0.96 | 7.51 | ± | 10.28 | <0.0001 |
| 9-HODE |  |  | 6.14 | ± | 3.03 | 225.74 | ± | 308.14 | <0.0001 |
| 13-HODE |  |  | 12.44 | ± | 5.72 | 263.98 | ± | 363.04 | <0.0001 |
| DHA | Anti-inflammatory | μM<br>(±SD) | 36.96 | ± | 17.63 | 39.67 | ± | 27.01 | 0.7135 |
| EPA |  |  | 37.26 | ± | 4.72 | 26.13 | ± | 21.59 | <0.001 |
| 17-HDHA |  | nM (±SD) | 1.88 | ± | 1.46 | 88.61 | ± | 107.3 | <0.0001 |
| 14-HDHA |  |  | 55.53 | ± | 54.4 | 93.11 | ± | 69.82 | 0.0006 |
| 18-HEPE |  |  | 0.49 | ± | 0.32 | 10.27 | ± | 14.16 | <0.0001 |
| RvD4 |  |  | 0.001* |  |  | 4.11 | ± | 3.94 | <0.0001 |
| Maresin 2 |  |  | 0.03 | ± | 0.01 | 0.42 | ± | 0.48 | <0.0001 |
| 5,6-EET |  |  | 0.24 | ± | 0.38 | 0.30 | ± | 0.29 | <0.0001 |
| 5,6-DHET |  |  | 0.38 | ± | 0.21 | 2.35 | ± | 1.73 | <0.0001 |
| 5,6-Ratio |  |  | 0.60 | ± | 0.72 | 0.21 | ± | 0.24 | <0.0001 |
| 8,9-EET |  |  | 0.21 | ± | 0.21 | 0.74 | ± | 0.88 | <0.0001 |
| 8,9-DHET |  |  | 0.22 | ± | 0.07 | 0.47 | ± | 0.32 | <0.0001 |
| 8,9-Ratio |  |  | 0.97 | ± | 0.86 | 2.63 | ± | 4.09 | 0.0098 |
| 11,12-EET |  |  | 0.56 | ± | 0.77 | 160.95 | ± | 122.47 | <0.0001 |
| 11,12-DHET |  |  | 0.53 | ± | 0.16 | 0.60 | ± | 0.32 | 0.315 |
| 11,12-Ratio |  |  | 1.09 | ± | 1.31 | 363.01 | ± | 407.70 | <0.0001 |
| 14,15-EET |  |  | 0.27 | ± | 0.35 | 0.43 | ± | 0.44 | <0.0001 |
| 14,15-DHET |  |  | 0.43 | ± | 0.14 | 0.52 | ± | 0.27 | 0.0672 |
| 14,15-Ratio |  |  | 0.68 | ± | 0.74 | 0.90 | ± | 0.73 | 0.0002 |

**Figure 3.**
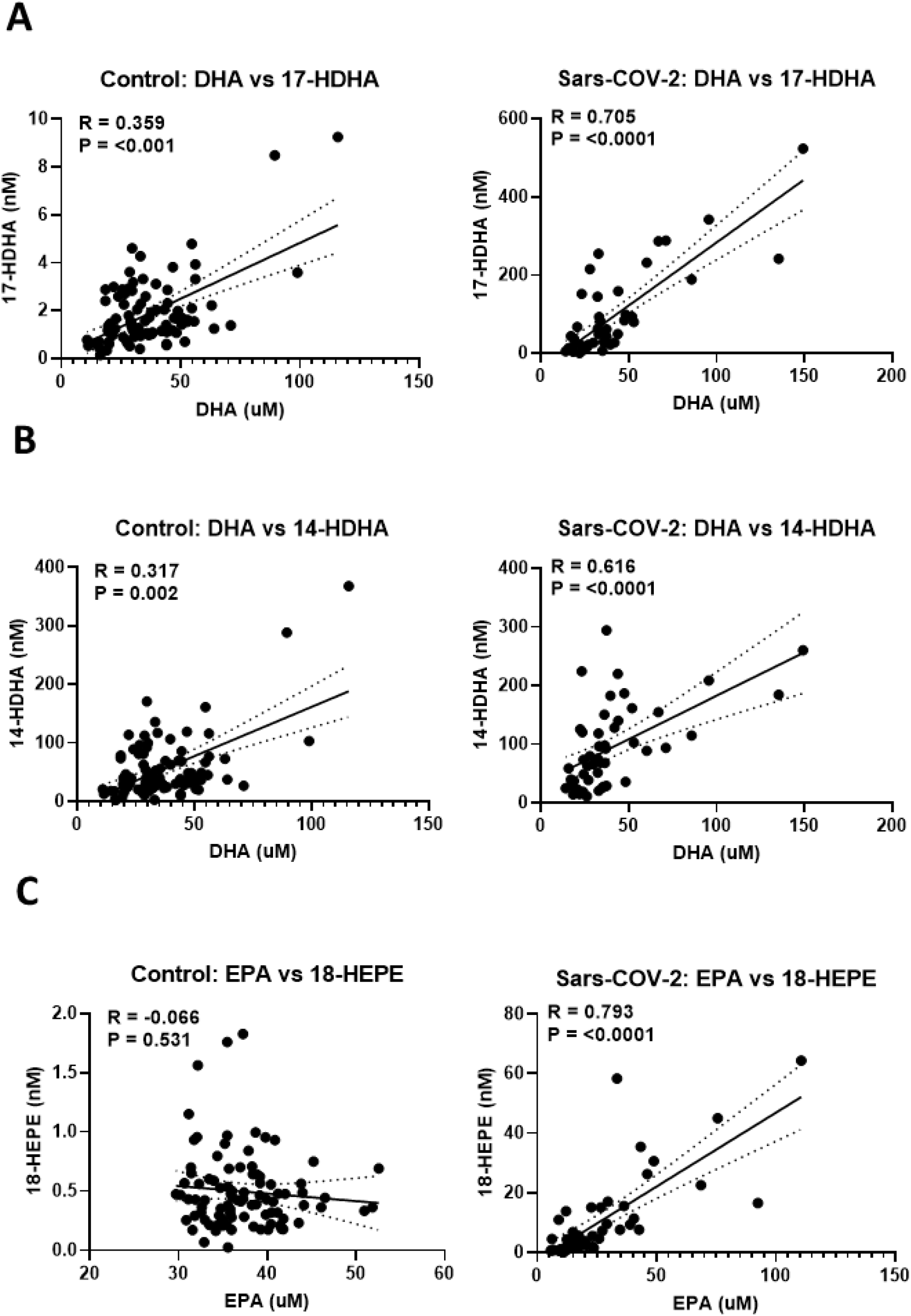
Correlation analysis of DHA & EPA and relevant down-stream SPM pathway metabolites (A) 17-HDHA, (B) 14-HDHA, and (C) 18-HEPE in SARS-CoV-2 (n=50) and age and sex controls (n=94). Data analysed by Spearman’s Rho.

**Figure 4.**
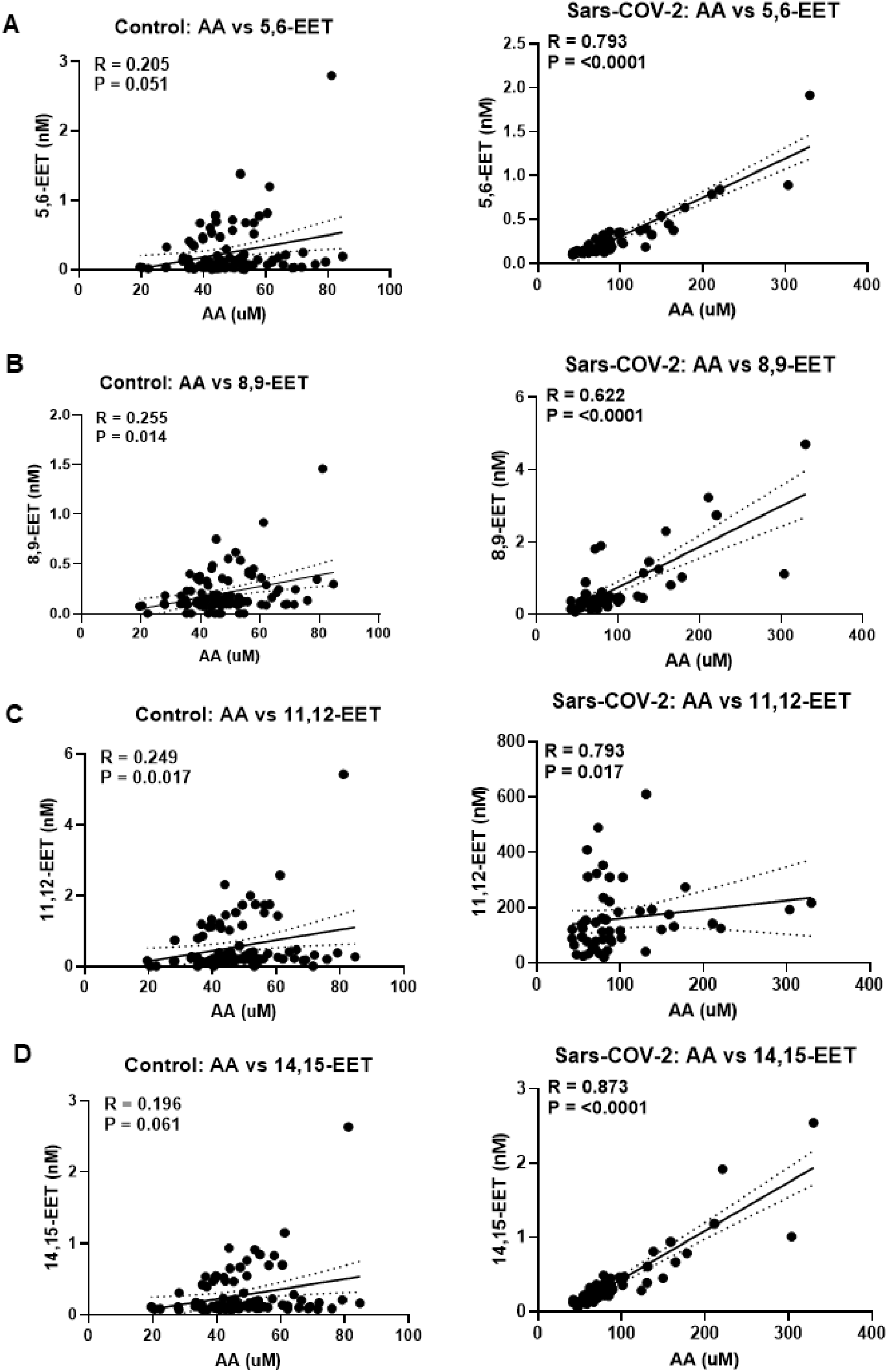
Correlation analysis between AA and down-stream CYP450 metabolites (A) 5,6-EET, (B) 8,9-EET, (C)1,12-EET, and (D) 14,15-EET in SARS-CoV-2 and control groups. Data analysed by Spearman’s Rho.

## Results

### Characteristics of SARS-COV-2 infection cohort and clinical features

Serum samples collected from hospital inpatients (22 female, 28 male) with confirmed diagnosis of Sars-CoV-2 (SARS-CoV-2) were studied (SI Table 2). 30 patients recovered and were discharged, 20 died, and 25 spent time in ICU during hospitalisation (SI Table 2).

**Table 2.** Correlation analysis between concentrations of bioactive lipid with anti-nucleocapsid & anti-spike antibody response in SARS-CoV-2 (n=50). Data analysed by Spearman’s Rho.

| Bioactive lipids | Correlation Analysis of Lipids vs Antibody binding |  |  |  |
| --- | --- | --- | --- | --- |
|  | Anti-Nucleocapsid |  | Anti-Spike |  |
|  | R Value | P Value | R Value | P Value |
| AA | 0.326 | **0.021** | 0.352 | **0.012** |
| LA | 0.249 | 0.081 | 0.382 | **0.006** |
| DHA | 0.369 | **0.009** | 0.337 | **0.017** |
| EPA | 0.419 | **0.002** | 0.540 | **<0.0001** |
| 17-HDHA | 0.269 | 0.059 | 0.537 | **<0.0001** |
| 18-HEPE | 0.366 | **0.009** | 0.410 | **0.003** |
| 14-HDHA | 0.434 | **0.002** | 0.293 | **0.039** |
| RvD4 | 0.211 | 0.142 | 0.449 | **0.001** |
| Maresin 2 | 0.282 | 0.055 | 0.343 | **0.018** |
| LXA4 | 0.051 | 0.723 | 0.324 | **0.022** |
| LXA5 | 0.222 | 0.133 | 0.436 | **0.002** |
| 5-HETE | 0.093 | 0.52 | 0.361 | **0.010** |
| 8-HETE | 0.404 | **0.004** | 0.419 | **0.002** |
| 9-HETE | 0.27 | 0.058 | 0.469 | **0.001** |
| 11-HETE | 0.279 | **0.05** | 0.464 | **0.001** |
| 12-HETE | 0.231 | 0.106 | 0.084 | 0.563 |
| 15-HETE | 0.259 | 0.069 | 0.407 | **0.003** |
| 16-HETE | 0.245 | 0.097 | 0.045 | 0.756 |
| 19-HETE | 0.138 | 0.344 | 0.314 | **0.028** |
| 20-HETE | 0.234 | 0.101 | 0.099 | 0.494 |
| TBxB2 | -0.26 | 0.071 | -0.052 | 0.722 |
| PGE2 | 0.049 | 0.744 | 0.275 | 0.056 |
| PGD2 | 0.113 | 0.445 | 0.283 | 0.052 |
| LTB4 | -0.041 | 0.776 | 0.326 | **0.021** |
| 5,6-EET | 0.357 | **0.011** | 0.271 | 0.057 |
| 5,6-DHET | 0.024 | 0.867 | 0.310 | **0.029** |
| 5,6-Ratio | 0.243 | 0.089 | -0.018 | 0.904 |
| 8,9-EET | 0.099 | 0.494 | 0.185 | 0.198 |
| 8,9-DHET | 0.063 | 0.663 | -0.104 | 0.474 |
| 8,9-Ratio | 0.064 | 0.664 | 0.164 | 0.257 |
| 11,12-EET | 0.239 | 0.093 | 0.084 | 0.564 |
| 11,12-DHET | 0.109 | 0.452 | -0.041 | 0.779 |
| 11,12-Ratio | 0.15 | 0.297 | 0.114 | 0.431 |
| 14,15-EET | 0.302 | **0.033** | 0.334 | **0.018** |
| 14,15-DHET | 0.023 | 0.874 | -0.194 | 0.178 |
| 14,15-Ratio | 0.339 | **0.016** | 0.443 | **0.001** |
| 13-oxo-ODE | 0.202 | 0.169 | 0.447 | **0.002** |
| 9-oxo-ODE | 0.243 | 0.154 | 0.482 | **0.003** |
| 9-HODE | 0.265 | 0.063 | 0.505 | **0.0002** |
| 13-HODE | 0.268 | 0.059 | 0.488 | **0.0003** |
| AEA | 0.017 | 0.907 | -0.216 | 0.131 |
| PEA | -0.071 | 0.626 | -0.083 | 0.567 |
| OEA | 0.063 | 0.666 | -0.060 | 0.679 |

### SARS-COV-2 serum has a distinct bioactive lipid profile

SARS-CoV-2 serum had high concentrations of ω-6 derived pro-inflammatory lipids (Table 1) and ω-6 and ω-3 derived anti-inflammatory SPM lipids (Table 1), which were significantly increased compared to age and sex matched control serum. In control serum, comparable to the healthy population, many of the anti-inflammatory SPM lipids were not detectable, or present at very low levels[29]. PLS-DA analysis of these lipids identified contributors to the separation between the SARS-CoV-2 and control serum (Figure 1A). 22 lipids from seven classes of lipids had a VIP score >1 and statistically underpinned the separation between the two clusters (SI Figure 4A). Although serum concentrations of the PUFAs (LA, EPA and DHA) were generally not different between the SARS-CoV-2 and control serum (Figure 1B, SI Table 1), concentrations of the down-stream bioactive lipids were markedly increased in SARS-CoV-2 serum (Figure 1B). There were substantially higher levels of the pro-inflammatory molecule PGD2, and the anti-inflammatory SPMs (17-HDHA, RvD4, LXA4, LXA5), endocannabinoids (2-AG, AEA) and, 11,12-EET in SARS-CoV-2 serum, compared to control serum (Table 1, SI Table 3). Our findings support a profound mobilisation of pro-resolving mediators following SARS-CoV-2 infection.

PLS-DA analysis revealed a clear separation between the SARS-CoV-2 and control serum for the three age groups (SI Figure 2A-C). Overall, the majority of the lipids contributing to the separation between the SARS-CoV-2 and control serum were the same for the three age groups studied (SI Figure 2D). This was confirmed by a univariate analysis (Figure 2, SI Table 4). Overall, the ability to mount a pro-resolution response was not markedly altered by age in our study.

Changes in the flux through enzymatic pathways generating SPMs may provide insight into novel treatments for Sars-CoV-2 infection. Serum levels of DHA and the downstream metabolites, 17-HDHA and 14-HDHA, were correlated in both SARS-CoV-2 and control serum (Figure 3A-B). However, levels of EPA and 18-HEPE were correlated in SARS-CoV-2 serum but not control serum, suggesting a potential upregulation of the E series resolving pathway following infection (Figure 3C).

Serum levels of AA were within a healthy range in the control group[30], but were elevated in SARS-CoV-2 serum across all age groups (SI Figure 5A, Table 1). Levels of the anti-inflammatory EETs and the downstream metabolites (DHETs) are presented both individually and as a ratio to reflect the activity of the sEH pathway (Table 1). The ratio of 11,12-EET:11,12-DHET was significantly increased in SARS-CoV-2 serum, compared to matched control serum (SI Figure 5B). There were some differences in the ratios of 8,9-EET:8,9-DHET and 14,15-EET:14,15-DHET but these were less consistent across the age groups for the two groups (Table 1). In the SARS-CoV-2 serum, levels of AA were correlated with all EETs, whereas in the control serum AA was only correlated with 8,9-EET and 11,12-EET (Figure 4B & 4C). These data suggest changes in the flux through the sEH pathway following SARS-CoV-2 infection, worthy of further future investigation.

**Figure 5.**
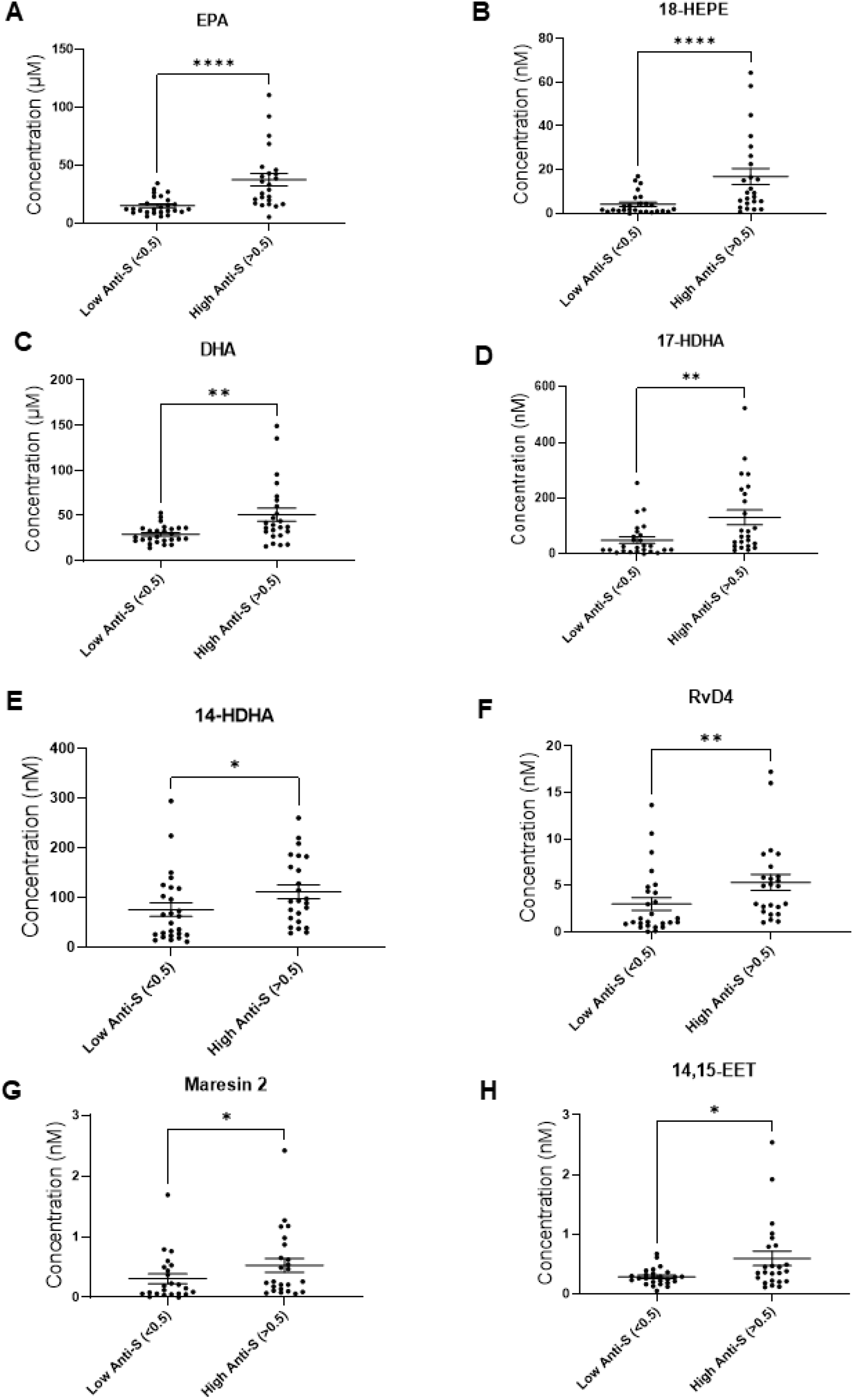
Serum concentration of significantly altered pro-resolution lipid mediators A) EPA, B) 18-HEPE, C) DHA, D) 17-HDHA, E) 14-HDHA, F) RvD4, G) Maresin 2, & H) 14,15-EET based on the anti-spike antibody response (low group <0.5, n=26, (high group>0.5, n=24) in SARS-CoV-2 patients. Groups were assessed for normal distribution using D’Agostino & Pearson test. Significance was assessed using Mann-Whitney test. *p=<0.05, **p=<0.01, ***p=<0.001, ****p=<0.0001.

### Associations between adaptive immune responses to SARS-COV-2 infection and serum levels of bioactive lipids

Anti-SARS-CoV-2 antibody binding to the nucleocapsid protein was interrogated using a reference antigen. Viral sequence data, where available, confirmed that the variants circulating in our cohort were either the Hu-1 strain, or the variant defining the 20B lineage (defined by the spike D614G variant), with no variability in nucleocapsid amino acid sequence[24]. There was a range of levels of anti-nucleocapsid (0.15 to 3.11 OD450) and anti-spike antibody (0.115-3.067 OD450) in the SARS-CoV-2 group. Anti-nucleocapsid data (Table 2) are discussed in SI. Based on the range of anti-spike antibody binding, patients were separated into two groups: low (<0.5, n=26) and high (>0.5, n=26), the subset of patients who died had significantly lower levels of anti-spike antibody binding (SI Figure 6). Levels of 18-HEPE, 17-HDHA, RvD4 and 14,15-EET were significantly higher in patients with an anti-spike antibody value >0.5 (Figure 5). There were statistically significant positive correlations between these lipids and anti-spike antibody binding for the entire group of patients (Table 2).

Analysis of serum levels of bioactive lipids early in the infection and the clinical outcome identified that levels of LA and 5,6-DHET were significantly lower in SARS-CoV-2 infected patients who died, and the ratio of 5,6-EET:5,6-DHET was higher in those patients that died compared to those that survived (SI Table 5).

## Discussion

SARS-CoV-2 infection was associated with robust increases in both ω-6 and ω-3 derived bioactive lipids which have well characterized roles in both pro- and anti-inflammatory responses[31]. Significant increases in serum levels of pro-inflammatory lipids included PGE2, TBXB2 and LTB4 and a range of bioactive lipids with well-established roles in dampening down / resolving inflammatory processes were evident in the SARS-CoV-2 group, compared to the age matched control group. Of particular note was the 47-fold increase in levels of the SPM precursor 17-HDHA and the SPMs RvD4 and Mar2, serum levels of which are normally close to, or below, the limits of detection in the healthy population[32]. Substantial increases in levels of some EETs and endocannabinoids also support a concerted anti-inflammatory response via multiple enzymatic pathways following SARS-CoV-2 infection. Our demonstration of a robust activation of the resolution pathways following SARS-CoV-2 infection, irrespective of patient age, points to a complex pathophysiological response which may be amenable to pharmacological intervention and provide new targets for treatment.

There was a broad range of anti-nucleocapsid and anti-spike responses in the SARS-CoV-2 group, indicative of adaptive immune response to infection. Consistent with a larger study, increased anti-spike responses were associated with improved clinical outcome[24]. SARS-CoV-2 patients with higher anti-spike responses (>0.5) had significantly increased levels of a number anti-inflammatory and resolution molecules as well as some pro-inflammatory lipids. The statistically altered bioactive lipids represent clusters of SPMs, lipoxins and EETs which either directly mediate resolution of inflammation or are metabolites in the resolution pathways and are generated by COX, LOX and CYP pathways. Levels of PUFA substrates were not substantially altered by SARS-CoV-2 infection or age, thus it is unlikely that substrate and therefore diet is major determining factor in the resolution response to SARS-CoV-2 infection. The strong correlations between PUFAs and their downstream SPM pathway metabolites in the SARS-CoV-2 infection group suggests that these enzymatic pathways are upregulated by this infection, particularly evident for the E series pathway. Our findings support future studies of the relationship between the antibody response to SARS-CoV-2 infection, activation of the resolution pathways and clinical outcome in a larger cohort of patients.

Our findings are consistent with the recent report of up-regulation of plasma and serum SPMs and increased expression of related enzymatic pathways in peripheral blood monocyte subsets in 19 patients infected with SARS-CoV-2[33] and findings by Archambault et al (2020)[34]. In addition, increased levels of pro-inflammatory bioactive lipids and anti-inflammatory SPMs, including RvD4, D5, D2, D1 and PDX have been reported in bronchoalveolar lavage from SARS-CoV-2 patient[35]. SPMs are already known to modulate acute lung injury and respiratory distress syndrome[36], supporting these findings following SARS-CoV-2 infection. Antibodies generated by B cells are critical to anti-viral immunity. The D series precursors and resolvins, including 17-HDHA, enhance human B cell antibody production by promoting differentiation towards an anti-body secreting phenotype[18]. In a pre-clinical murine model of influenza immunization, 17-HDHA treatment increased antigen specific antibody responses and protected against live influenza virus infection[17]. These data suggest that a robust generation of 17-HDHA following infection not only acts to counter pro-inflammatory responses but may also facilitate the response of B cells to mount an antibody response. To date there are no studies of the effects of the SPMs on SARS-CoV-2 infection in patients, however, it has been reported that both RvD1 and RvD2 have beneficial effects on inflammatory responses in SARS-CoV-2 infected macrophages[37].

Current evidence suggests that treatments for SARS-CoV-2 infection, alongside vaccinations, will remain a priority in the immediate future. Identification of the critical steps involved in the up-regulation of these anti-inflammatory pathways may be instructive for the development of interventions aimed at dampening the inflammatory response or promoting the clearance of the inflammation arising from SARS-CoV-2 infection. Dexamethasone has been identified as an important treatment to promote recovery from SARS-CoV-2 infection. Dexamethasone has been shown to increase SPM levels in a small number of SARS-CoV-2 infected patients[33], healthy volunteers[38] and in allergic airway inflammation[39]. Future investigation of the potential contribution of SPMs to the beneficial effects of dexamethasone in patients with SARS-CoV-2 infection in a larger cohort will improve further our understanding of the potential mechanisms of action of this treatment.

There are a number of study limitations. Serum samples were collected from patients hospitalised with SARS-CoV-2 for clinical diagnostic tests during the first wave of the pandemic in the UK. Due to the clinical pressures within the system at the time some clinical data, such as BMI, was not collected. It is known that levels of SPMs are decreased with increased BMI, which could potentially contribute to SARS-CoV-2 related morbidities and mortalities[40]. Although we have data relating to whether a patient was admitted to intensive care, clinical decision making was related to multiple factors beyond the severity of the infection and therefore further analysis of potential impact has not been performed. Serum samples were collected within the first few days of hospital admission and represent a snapshot of the anti-nucleocapsid and anti-spike response and the lipid levels at a point in time. It is important to note that levels of antibodies change over time, however we do not have matched longitudinal data. The anti-nucleocapsid and anti-spike signal provides an indication of the potency of the adaptive immune response following infection. The viral genome sequencing data available for these patients indicated that the nucleocapsid amino acid sequence was completely conserved between infections. While it is possible that there may be mismatches between the antigen used in our assay and the strain of infecting virus that mean that antibodies are present which are undetected by our assay, at the time of sampling there was minimal genetic diversity in UK isolates[www.nextrstrain.org]. To mitigate against this, we used ELISAs against two antigens.

In summary, our findings highlight that SARS-CoV-2 infection can lead to very robust activation of the pathways that generate the specialised pro-resolving molecules and other anti-inflammatory bioactive lipids. This new knowledge supports the future investigation of these pathways which may inform the development of novel anti-inflammatory treatments for SARS-CoV-2 infection. Furthermore, these new datasets provide us opportunities to explore further the underlying molecular pathways that regulate the resolution pathways in humans, with the goal of identifying novel approaches for the development of new therapeutics for other infections and chronic inflammatory diseases.

## Data Availability

All data produced in the present study are available upon reasonable request to the authors

## Notes

### Author Contributions

VC, DAB, DHK, & AWT conceived the study, secured the funding, supervised data collection and analysis, and prepared manuscript.

JT, RJ, & CO performed lipidomic analysis of samples.

AWT, PJT, & WLI supplied clinical samples, performed measurement of anti-nucleocapsid and prepared manuscript.

AMV & SAG provided control serum samples.

JT & RJ performed statistical analysis and prepared manuscript.

EL provided clinical insight into data interpretation and prepared manuscript.

All authors reviewed manuscript and had final responsibility for the decision to submit for publication.

## Acknowledgements

The authors thank Professors David Walsh and John Gladman for their advice in preparation of the manuscript. The authors also thank Dr Divyateja Hrushikesh and colleagues from the Queens Medical Centre for providing samples and clinical data.

## Disclaimer

The source of funding had no influence over the design or conduct of the study.

