## Supplementary Materials for "Serum levels of specialised pro-resolving molecule pathways are greatly increased in SARS-CoV-2 patients and correlate with markers of the adaptive immune response"

### Supplementary Information

### Supplementary Methods

#### LC-MS/MS lipidomic profiling

Samples were stored at -80°C before analysis and thawed on ice prior to extraction. 400 µL of calibration standards, QC samples, and study samples (available volumes of study samples were measured and recorded if less than 400 µL for post analysis calculations) were spiked with 40 µL of an internal standard solution containing: AA-d8 (1.7 µM), PGD2-d4 (0.2 µM), and 15-HETE-d8 (1.3 µM), RvD2-d4 (0.2 µM), and AEA-d8 (0.8 µM). Bioactive lipids were isolated by solid phase extraction using a C18 cartridge.

The HPLC system used was ExionLC™ Series UHPLC (Applied Biosystem, Foster City, CA, USA). The HPLC Column used was Acquity UPLC BEH C18 1.7 µm (150 x 2.1 mm) with guard column (Acquity UPLC BEH C18 1.7 µm, VanGuard Pre-Column 3/Pk, 2.1 x 5 mm). Mobile phase A was 0.02% formic acid in 100% water; mobile phase B was formic acid in methanol/acetonitrile (1:4, v/v) with a flow rate of 0.3 mL/min.

The MS system used was an Applied Biosystem MDS SCIEX 6500 Q-Trap hybrid triple-quadrupole–linear ion trap mass spectrometer (Applied Biosystem, Foster City, CA, USA) equipped with an electrospray ionisation (ESI) interface.

Quantification of each analyte was calculated using a fully extracted calibration standard curve. Normalisation was performed against the aliquoted sample volume to produce final concentration of each individual lipid present in serum samples. Quantification was performed using MultiQuant V.3.0.3 software. Identification of each compound in plasma samples was confirmed by LC retention times, corresponding secondary product ion transition, and presence in a solution of disambiguated analytes. The peak area ratio of each analyte was used to calculate the molar concentration using the calibration standard curve.

Each measured lipid had its identity confirmed using the following criteria: (1) presence of a peak with S/N ratio of greater than 5, (2) presence of reference standard spiked QCs and calibration line, (3) matching retention time with authentic reference standards, (4) ≥10 data points per peak, and (4) matching of at least 2 diagnostic fragment ions from a reference standard.

To assess any effect of the SARS-CoV-2 deactivation procedure on serum levels of lipid mediators, healthy serum from young volunteers (n=10) was used to replicate the sample storage and handling of the control and SARS-CoV-2 study samples. Serum was pooled to prepare n=12 identical QC samples. To replicate control group sample handling, n=6 of these samples were immediately stored at -80°C. To mimic the sample handling for the SARS-CoV-2 samples, n=6 QC samples were stored at 4°C for 24 h and then at RT for 4 h with 1% triton-X 100. All samples were extracted and analysed by LC-MS/MS simultaneously and the levels of lipid mediators quantified and compared.

### Supplementary Results

#### Associations between adaptive immune responses to SARS-COV-2 infection and serum levels of bioactive lipids

Based on the ranges, patients were divided into three groups of anti-nucleocapsid response (<0.5 (n=16), 0-5-2.5 (n=11) and >2.5 (n=22) OD450) and levels of bioactive lipids between the groups compared.  Serum levels of 14-HDHA, 11,12 EET and 8, 12, and 20-HETE were significantly higher in those samples with an anti-nucleocapsid response >2.5 (Table 4). Serum levels of AA, 8-HETE, DHA, EPA, 14-HDHA and 18-HEPE were positively correlated with anti-nucleocapsid levels (Table 4).

### Supplemntary Figures

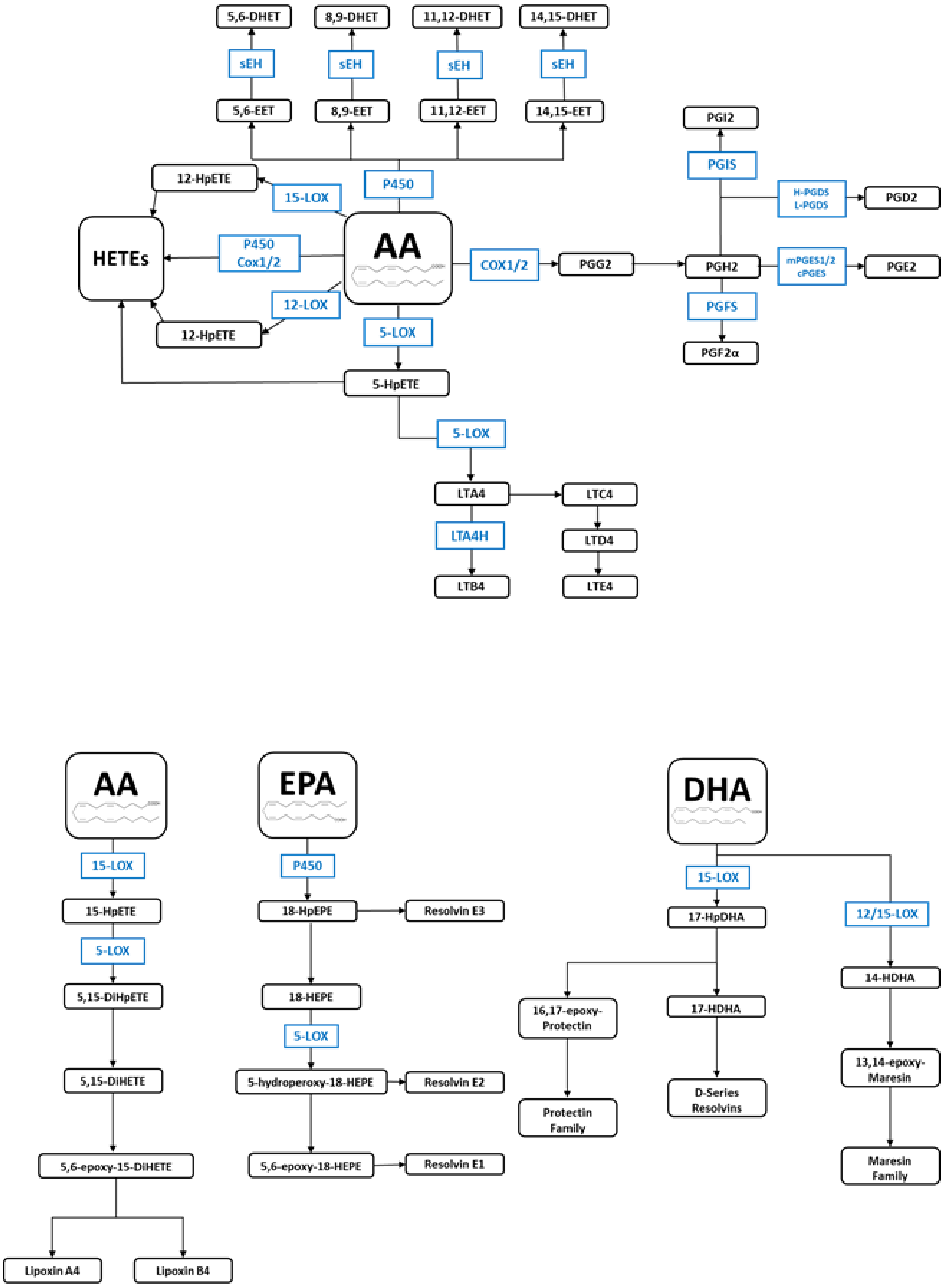


SI Figure 1. Biosynthetic pathways of metabolites from the arachidonic acid, eicosapentaenoic acid, and docosahexaenoic acid cascades.

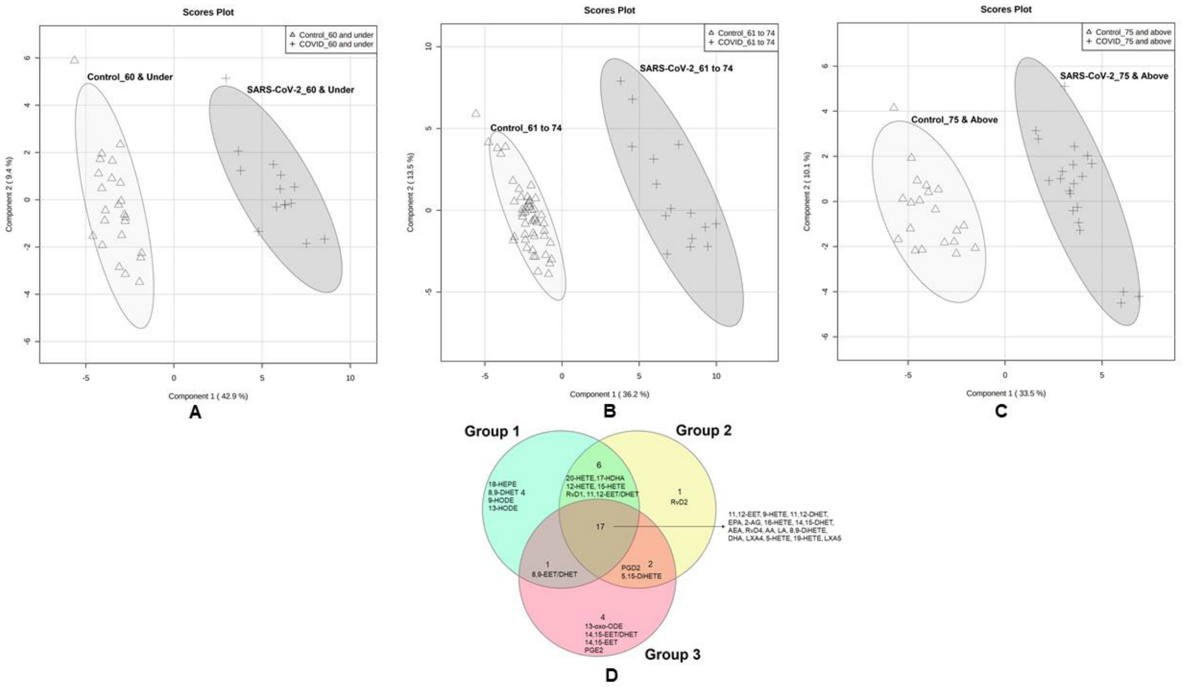


SI Figure 2**.** Partial least square discrimination analysis (PLS-DA) for the 44 serum lipids quantified in SARS-CoV-2 (n=50) and age and sex matched controls (n=94), stratified by age. A: Group 1 (60 and under) with R2 = 0.966, Q2 = 0.941 and Accuracy = 1.0; B: Group 2 (61 to 74) with R2 = 0.986, Q2 = 0.969 and Accuracy = 1.0; C: Group 3 (75 and above) with R2 = 0.952, Q2 = 0.928 and Accuracy = 1.0. D: Venn diagram showing the common versus unique lipid mediators obtained from PLS-DA analysis (VIP>1) of the age stratified samples.

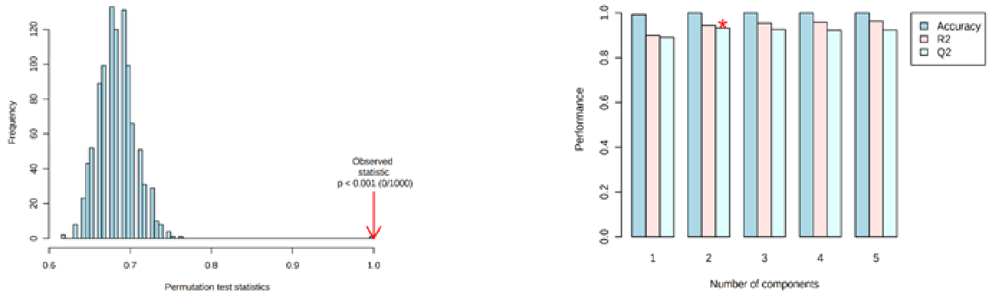

A. Control Vs SARS-CoV-2

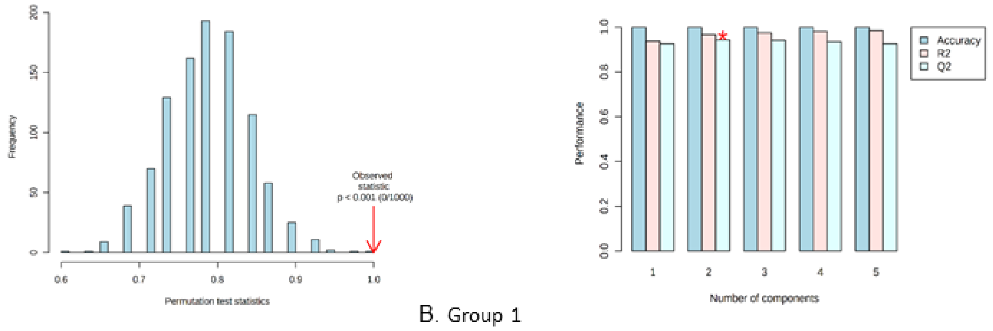

B. Group 1

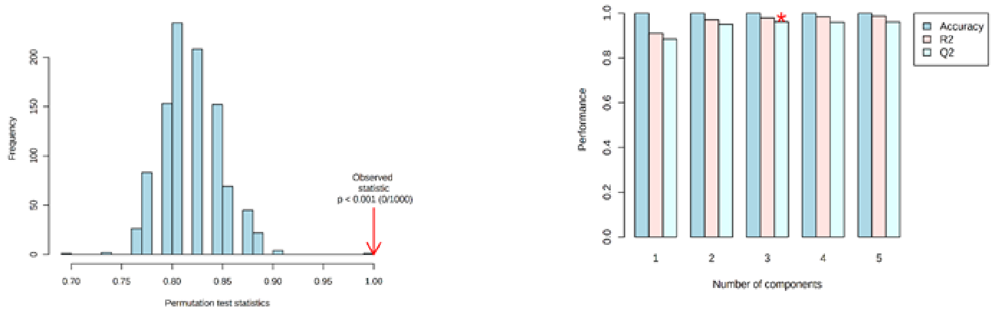

C. Group 2

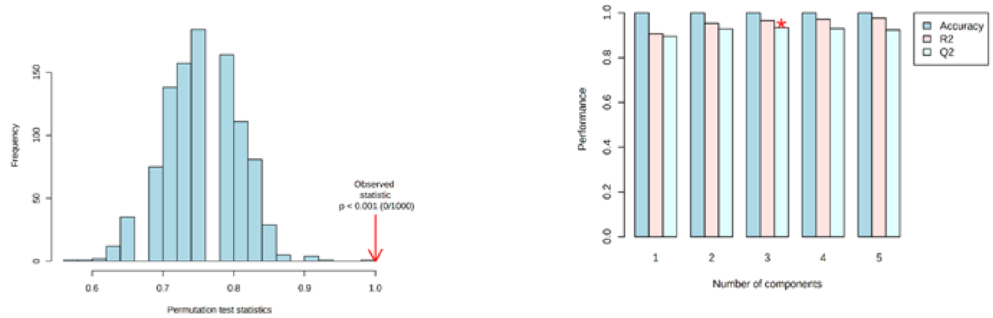

D. Group 3

SI Figure 3. Permutation tests and cross validation for PLS-DA model of A) Control Vs SARS-CoV-2, B) Group 1 (Control_60 & Under Vs SARS-CoV-2_60 & Under), C) Group 2 (Control_61 to 74 Vs SARS-CoV-2_61 to 74) and D) Group 3 (Control_75 & Above Vs SARS-CoV-2_75 & Above).

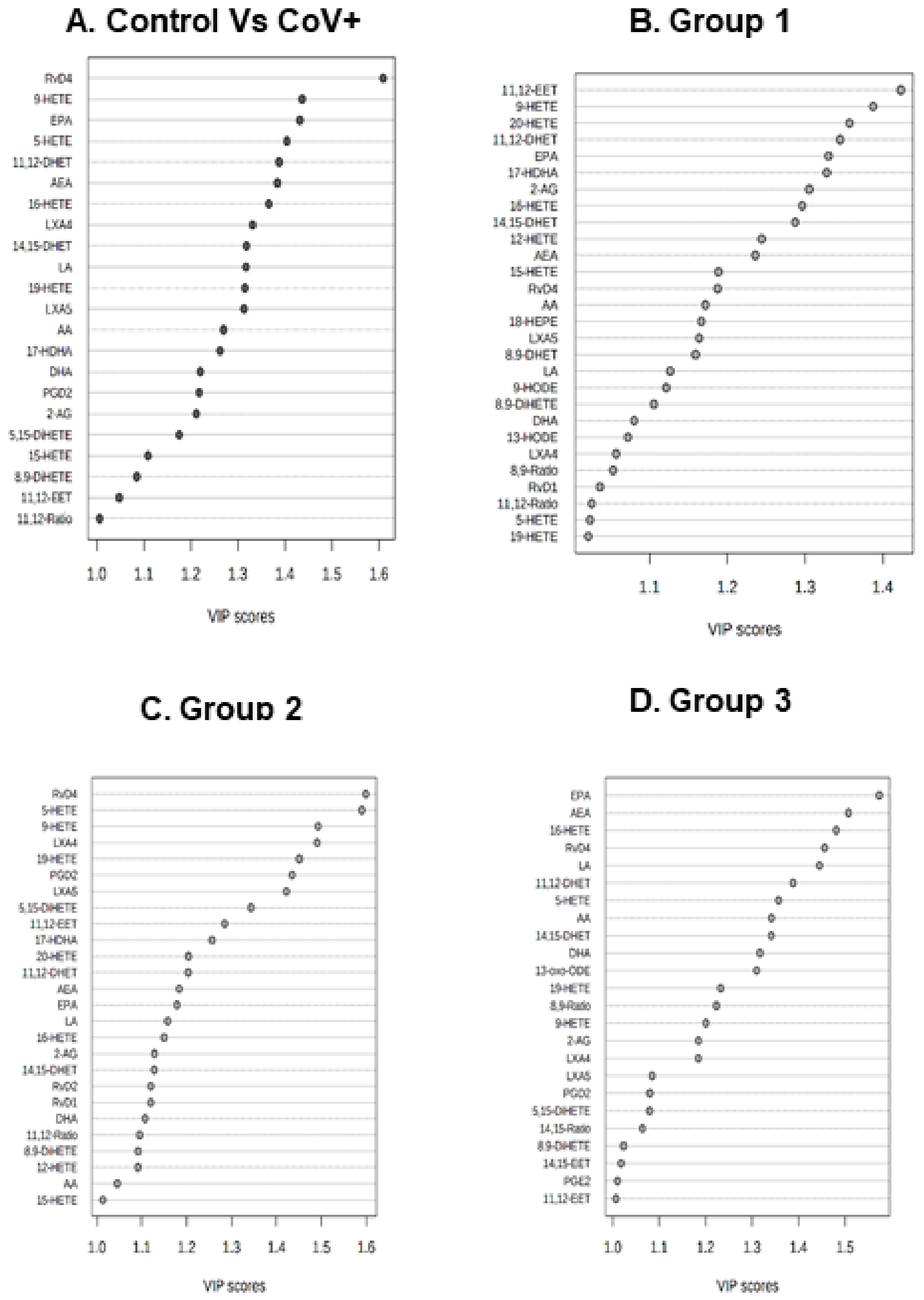


SI Figure 4. VIP graphs showing important discriminatory lipids (VIP>1) in A) Control Vs SARS-CoV-2, B) Group 1 (Control_60 & Under Vs SARS-CoV-2_60 & Under), C) Group 2 (Control_61 to 74 Vs SARS-CoV-2_61 to 74) and D) Group 3 (Control_75 & Above Vs SARS-CoV-2_75 & Above).

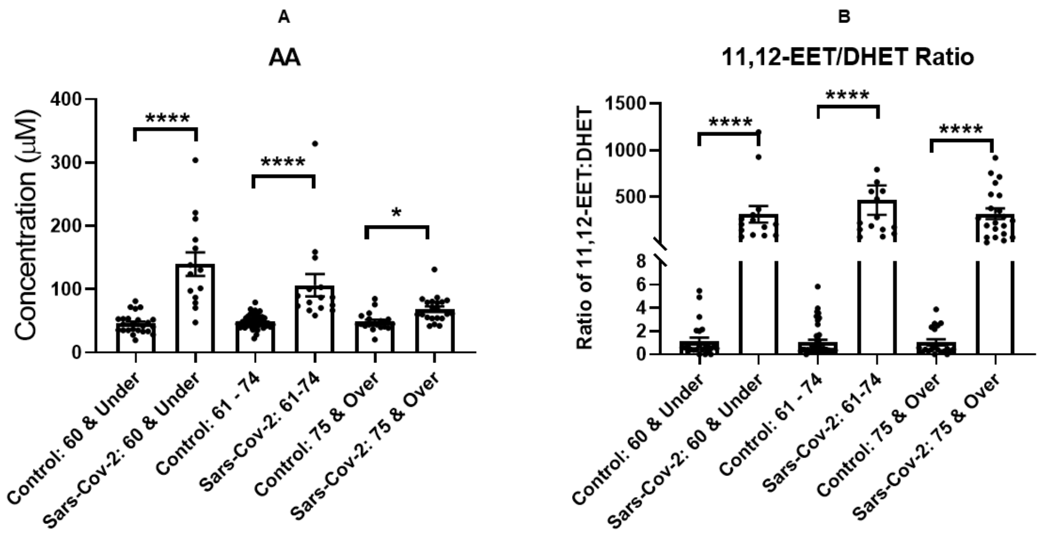


SI Figure 5**.** Serum concentrations of (A) AA (B) the ratio of 11,12-EET: DHET in SARS-CoV-2 (n=50) and age and sex matched controls (n=94) stratified by age group. Groups were assessed for normal distribution using D'Agostino & Pearson test. Significance was assessed using Kruskal-Wallis test correcting for multiple comparisons using Dunn's test. *p=<0.05, **p=<0.01, ***p=<0.001, ****p=<0.0001.

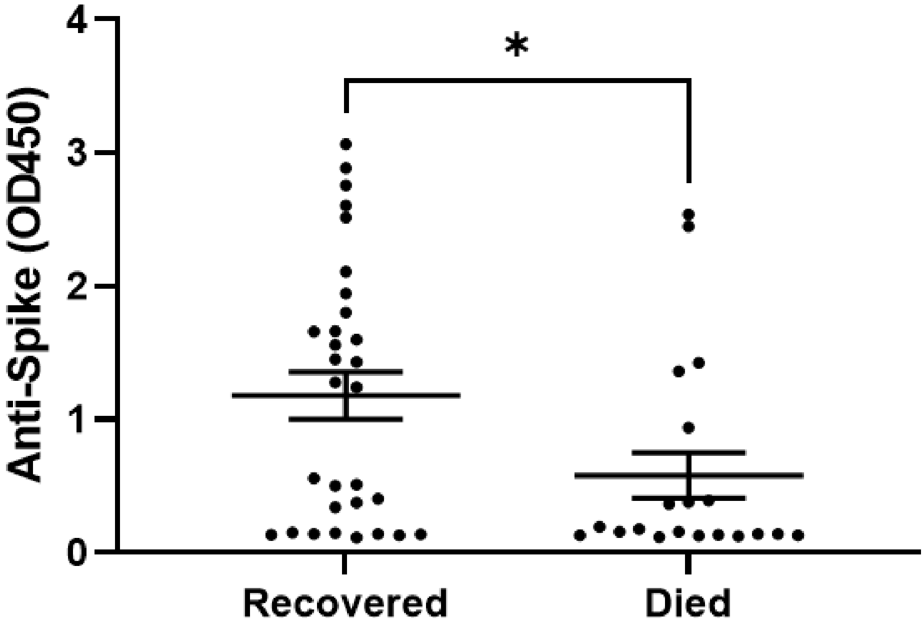


SI Figure 6. Levels of anti-spike antibody binding in SARS-CoV-2 patients stratified by clinical outcome, recovered (n=30) and died (n=20). Statistical analysis by Mann-Whitney test showed lower levels of anti-spike antibody binding in those patients who died (p=0.0133).

**Supplemntary Tables**

SI Table 1. List of key lipid markers involved in discriminatory profile of SARS-CoV-2 samples based on VIP scores (>1) along with false discovery rate (FDR) in age stratified groups.

| Group 1 | | | Group 2 | | | Group 3 | | |
|---|---|---|---|---|---|---|---|---|
| **Lipids** | **VIP** | **FDR** | **Lipids** | **VIP** | **FDR** | **Lipids** | **VIP** | **FDR** |
| 11,12-EET | 1.42 | 8.5E-13 | RvD4 | 1.60 | 4.0E-23 | EPA | 1.57 | 2.4E-11 |
| 9-HETE | 1.38 | 1.3E-11 | 5-HETE | 1.59 | 1.0E-22 | AEA | 1.51 | 7.8E-10 |
| 20-HETE | 1.35 | 1.0E-10 | 9-HETE | 1.49 | 2.7E-17 | 16-HETE | 1.48 | 2.2E-09 |
| 11,12-DHET | 1.34 | 1.9E-10 | LXA4 | 1.49 | 2.7E-17 | RvD4 | 1.46 | 5.5E-09 |
| EPA | 1.33 | 3.9E-10 | 19-HETE | 1.45 | 1.0E-15 | LA | 1.45 | 7.4E-09 |

| | | | | | | | | |
|---|---|---|---|---|---|---|---|---|
| 17-HDHA | 1.32 | 3.9E-10 | PGD2 | 1.43 | 3.5E-15 | 11,12-DHET | 1.39 | 6.9E-08 |
| 2-AG | 1.30 | 1.4E-09 | LXA5 | 1.42 | 8.8E-15 | 5-HETE | 1.36 | 1.9E-07 |
| 16-HETE | 1.30 | 2.0E-09 | 5,15-DiHETE | 1.34 | 2.2E-12 | AA | 1.34 | 2.6E-07 |
| 14,15-DHET | 1.29 | 3.0E-09 | 11,12-EET | 1.28 | 6.4E-11 | 14,15-DHET | 1.34 | 2.6E-07 |
| 12-HETE | 1.24 | 2.5E-08 | 17-HDHA | 1.26 | 2.6E-10 | DHA | 1.32 | 5.2E-07 |
| AEA | 1.24 | 3.3E-08 | 20-HETE | 1.20 | 2.8E-09 | 13-oXo-ODE | 1.31 | 6.0E-07 |
| 15-HETE | 1.19 | 2.2E-07 | 11,12-DHET | 1.20 | 2.8E-09 | 19-HETE | 1.23 | 5.5E-06 |
| RvD4 | 1.19 | 2.2E-07 | AEA | 1.18 | 6.1E-09 | 8,9-RATIO | 1.22 | 6.5E-06 |
| AA | 1.17 | 3.8E-07 | EPA | 1.18 | 7.1E-09 | 9-HETE | 1.20 | 1.1E-05 |
| 18-HEPE | 1.17 | 4.4E-07 | LA | 1.16 | 1.6E-08 | 2-AG | 1.18 | 1.4E-05 |
| LXA5 | 1.16 | 4.6E-07 | 16-HETE | 1.15 | 2.0E-08 | LXA4 | 1.18 | 1.4E-05 |
| 8,9-DHET | 1.16 | 5.1E-07 | 2-AG | 1.13 | 4.3E-08 | LXA5 | 1.08 | 1.2E-04 |
| LA | 1.13 | 1.5E-06 | 14,15-DHET | 1.13 | 4.3E-08 | PGD2 | 1.08 | 1.2E-04 |
| 9-HODE | 1.12 | 1.7E-06 | RvD2 | 1.12 | 5.2E-08 | 5,15-DiHETE | 1.08 | 1.2E-04 |
| 8,9-DiHETE | 1.11 | 2.7E-06 | RvD1 | 1.12 | 5.2E-08 | 14,15-Ratio | 1.06 | 1.6E-04 |
| DHA | 1.08 | 5.6E-06 | DHA | 1.11 | 8.0E-08 | 8,9-DiHETE | 1.02 | 3.3E-04 |
| 13-HODE | 1.07 | 6.8E-06 | 11,12-Ratio | 1.10 | 1.2E-07 | 14,15-EET | 1.02 | 3.5E-04 |
| LXA4 | 1.06 | 1.0E-05 | 8,9-DiHETE | 1.09 | 1.3E-07 | PGE2 | 1.01 | 3.8E-04 |
| 8,9-Ratio | 1.05 | 1.1E-05 | 12-HETE | 1.09 | 1.3E-07 | 11,12-EET | 1.01 | 3.9E-04 |
| RvD1 | 1.04 | 1.6E-05 | AA | 1.05 | 5.7E-07 | | | |
| 11,12-Ratio | 1.03 | 2.1E-05 | 15-HETE | 1.01 | 1.5E-06 | | | |
| 5-HETE | 1.02 | 2.2E-05 | | | | | | |
| 19-HETE | 1.02 | 2.2E-05 | | | | | | |

SI Table 2**.** Clinical characteristics of SARS-CoV-2 and control groups.

| **Characteristics** | **Control** | **Sars-CoV-2** |
|---|---|---|
| Total | 91 | 50 |
| Sex (M/F) | 30/61 | 28/22 |
| Age (Mean, Range) | 67 (45-85) | 69.3 (35-91) |
| Days between diagnosis & sample collection (Mean, Range) | - | 7 (0-21) |
| Recovered/Died | - | 30/20 |
| no-ICU/ICU | - | 25/25 |
| CRP (Mean, Range) | - | 172.9 (11-489) |
| Anti-nucleocapsid (Mean, Range) | - | 1.81 (0.12-3.1) |
| Anti-Spike (Mean, Range) | - | 0.939 (0.115-3.067) |

SI Table 3. Calculated concentrations of selected bioactive lipids quantified in SARS-CoV-2 (n=50) and age and sex matched controls (n=94), and stratified by age group.

| Lipid | | Healthy – Under 30 | Control – all groups | Control – 60 & Under | Control – 61-74 | Control – 75 & Over | Sars-COV-2 – all groups | Sars-COV-2 – 60 & Under | Sars-COV-2 – 61-74 | Sars-COV-2 – 75 & Over |
|---|---|---|---|---|---|---|---|---|---|---|
| DHA | µM ± SD | 259 ± 202.9 | 36.96 ± 17.63 | 29.36 ± 14.3 | 37.27 ± 15.06 | 45.37 ± 22.35 | 39.67 ± 27.01 | 53.23 ± 53.23 | 44.74 ± 31.39 | 27.02 ± 8.6 |
| EPA | | 42.38 ± 54.33 | 37.26 ± 4.72 | 38 ± 5.28 | 36.48 ± 4.28 | 38.24 ± 4.69 | 26.13 ± 21.59 | 38.13 ± 38.13 | 31.15 ± 24.24 | 14.55 ± 7.91 |
| AA | | 127.3 ± 41.64 | 47.98 ± 12.75 | 45.69 ± 15.3 | 48.88 ± 10.73 | 48.57 ± 13.53 | 99.61 ± 60.21 | 139.5 ± 139.5 | 106.3 ± 65.98 | 68.25 ± 19.79 |
| LA | | 18.29 ± 8.64 | 307.8 ± 109.7 | 271.9 ± 110 | 305.2 ± 102.4 | 357 ± 107.5 | 213.8 ± 108.4 | 314.1 ± 314.1 | 197.2 ± 81.98 | 158.8 ± 46.51 |
| 17-HDHA | nM ± SD | 1.92 ± 0.14 | 1.88 ± 1.46 | 1.33 ± 0.83 | 1.82 ± 0.98 | 2.69 ± 2.37 | 88.61 ± 107.3 | 124.4 ± 124.4 | 120.9 ± 143.6 | 41.66 ± 57.29 |
| 14-HDHA | | 2.23 ± 1.4 | 55.53 ± 54.4 | 35.6 ± 28.3 | 52.48 ± 34.86 | 86.77 ± 90.99 | 93.11 ± 69.82 | 95.91 ± 95.91 | 99.29 ± 66.16 | 86.83 ± 78.72 |
| 18-HEPE | | 0.25 ± 0.15 | 0.49 ± 0.32 | 0.34 ± 0.18 | 0.5 ± 0.28 | 0.67 ± 0.44 | 10.27 ± 14.16 | 14.52 ± 14.52 | 16.07 ± 19.94 | 3.29 ± 3.45 |
| RvD4 | | Not Detected | Not Detected | Not Detected | Not Detected | Not Detected | 4.11 ± 3.94 | 4.9 ± 4.9 | 5.26 ± 5.08 | 2.76 ± 3.36 |
| Maresin 2 | | Not Detected | 0.03 ± 0.01 | 0.02 ± 0.02 | 0.03 ± 0.01 | 0.03 ± 0.02 | 0.42 ± 0.48 | 0.5 ± 0.5 | 0.6 ± 0.63 | 0.25 ± 0.38 |
| 5,6-EET | | 2.63 ± 1.58 | 0.24 ± 0.38 | 0.32 ± 0.59 | 0.22 ± 0.27 | 0.21 ± 0.24 | 0.30 ± 0.29 | 0.41 ± 0.41 | 0.36 ± 0.43 | 0.19 ± 0.07 |
| 5,6-DHET | | 0.19 ± 0.06 | 0.38 ± 0.21 | 0.42 ± 0.31 | 0.38 ± 0.16 | 0.32 ± 0.14 | 2.35 ± 1.73 | 1.86 ± 1.86 | 1.79 ± 0.9 | 3.09 ± 2.14 |
| 5,6-Ratio | | 13.99 ± 8 | 0.60 ± 0.72 | 0.68 ± 0.92 | 0.52 ± 0.56 | 0.67 ± 0.77 | 0.21 ± 0.24 | 0.34 ± 0.34 | 0.25 ± 0.21 | 0.1 ± 0.09 |
| 8,9-EET | | 0.26 ± 0.09 | 0.21 ± 0.21 | 0.23 ± 0.29 | 0.19 ± 0.18 | 0.21 ± 0.18 | 0.74 ± 0.88 | 0.94 ± 0.94 | 0.89 ± 1.2 | 0.51 ± 0.37 |
| 8,9-DHET | | 0.19 ± 0.02 | 0.22 ± 0.07 | 0.22 ± 0.09 | 0.23 ± 0.07 | 0.19 ± 0.06 | 0.47 ± 0.32 | 0.36 ± 0.36 | 0.41 ± 0.37 | 0.58 ± 0.3 |
| 8,9-Ratio | | 1.3 ± 0.37 | 0.97 ± 0.86 | 0.97 ± 0.7 | 0.88 ± 0.82 | 1.18 ± 1.07 | 2.63 ± 4.09 | 3.45 ± 3.45 | 3.77 ± 5.77 | 1.3 ± 1.45 |
| 11,12-EET | | 6.41 ± 4.52 | 0.56 ± 0.77 | 0.66 ± 1.13 | 0.54 ± 0.61 | 0.49 ± 0.53 | 160.95 ± 122.47 | 139.7 ± 139.7 | 160.4 ± 120.9 | 175.5 ± 148.7 |
| 11,12-DHET | | 0.45 ± 0.12 | 0.53 ± 0.16 | 0.51 ± 0.15 | 0.55 ± 0.17 | 0.51 ± 0.14 | 0.60 ± 0.32 | 0.63 ± 0.63 | 0.51 ± 0.28 | 0.65 ± 0.36 |
| 11,12-Ratio | | 14.12 ± 9.81 | 1.09 ± 1.31 | 1.15 ± 1.45 | 1.08 ± 1.31 | 1.05 ± 1.14 | 363.01 ± 407.70 | 314.6 ± 314.6 | 466.8 ± 591.4 | 321.2 ± 257.5 |

| | | | | | | | | | | |
|---|---|---|---|---|---|---|---|---|---|---|
| 14,15-EET | | 2.14 ± 1.03 | 0.27 ± 0.35 | 0.33 ± 0.53 | 0.26 ± 0.26 | 0.24 ± 0.22 | 0.43 ± 0.44 | 0.65 ± 0.65 | 0.49 ± 0.58 | 0.26 ± 0.11 |
| 14,15-DHET | | 0.33 ± 0.1 | 0.43 ± 0.14 | 0.4 ± 0.14 | 0.45 ± 0.14 | 0.41 ± 0.12 | 0.52 ± 0.27 | 0.5 ± 0.5 | 0.5 ± 0.33 | 0.55 ± 0.27 |
| 14,15-Ratio | | 6.5 ± 2.9 | 0.68 ± 0.74 | 0.74 ± 0.82 | 0.66 ± 0.74 | 0.65 ± 0.64 | 0.90 ± 0.73 | 1.37 ± 1.37 | 0.99 ± 0.66 | 0.52 ± 0.27 |

SI Table 4. Adjusted P values for Dunn's multiple comparison test comparing lipid levels between age groups in control and SARS-CoV-2 cohorts.

| **Lipid** | **Control: 60 & Under vs. Control: 61 - 74** | **Control: 60 & Under vs. Control: 75 & Over** | **Control: 61 - 74 vs. Control: 75 & Over** | **Sars-COV-2: 60 & Under vs. Sars-COV-2: 61-74** | **Sars-COV-2: 60 & Under vs. Sars-COV-2: 75 & Over** | **Sars-COV-2: 61-74 vs. Sars-COV-2: 75 & Over** | **Control: 60 & Under vs. Sars-COV-2: 60 & Under** | **Control: 61 - 74 vs. Sars-COV-2: 61-74** | **Control: 75 & Over vs. Sars-COV-2: 75 & Over** |
|---|---|---|---|---|---|---|---|---|---|
| 5,6-EET | ns | ns | ns | ns | ns | ns | 0.0266 | ns | ns |
| 5,6-DHET | ns | ns | ns | ns | ns | ns | 0.0002 | <0.0001 | <0.0001 |
| 5,6-Ratio | ns | ns | ns | ns | ns | ns | ns | ns | 0.0007 |
| 8,9-EET | ns | ns | ns | ns | ns | ns | 0.0027 | 0.0027 | 0.0048 |
| 8,9-DHET | ns | ns | ns | ns | ns | ns | ns | ns | <0.0001 |
| 8,9-Ratio | ns | ns | ns | ns | ns | ns | ns | 0.1597 | ns |
| 11,12-EET | ns | ns | ns | ns | ns | ns | <0.0001 | <0.0001 | <0.0001 |
| 11,12-DHET | ns | ns | ns | ns | ns | ns | ns | ns | ns |
| 11,12-Ratio | ns | ns | ns | ns | ns | ns | <0.0001 | <0.0001 | <0.0001 |
| 14,15-EET | ns | ns | ns | ns | ns | ns | 0.0145 | 0.1221 | ns |
| 14,15-DHET | ns | ns | ns | ns | ns | ns | ns | ns | ns |
| 14,15-Ratio | ns | ns | ns | ns | ns | ns | ns | 0.0456 | ns |
| EPA | ns | ns | 0.0071 | ns | ns | ns | ns | ns | <0.0001 |
| DHA | ns | ns | 0.0336 | ns | ns | 0.011 | 0.0298 | ns | 0.0116 |
| 17-HDHA | ns | ns | ns | ns | ns | ns | <0.0001 | <0.0001 | 0.0004 |
| 14-HDHA | ns | ns | ns | ns | ns | ns | 0.0029 | ns | ns |
| 18-HEPE | ns | ns | ns | ns | ns | ns | <0.0001 | <0.0001 | 0.0132 |

| | | | | | | | | | |
|---|---|---|---|---|---|---|---|---|---|
| Maresin 2 | ns | ns | ns | ns | ns | ns | <0.0001 | <0.0001 | 0.0045 |
| RvD4 | - | - | - | ns | ns | ns | - | - | - |

SI Table 5. Concentrations of bioactive lipids quantified in Sars-CoV-2 (n=50) patients who recovered from infection (n=30) versus who died from infection (n=20). Statistical analysis by Mann-Whitney Test

| **Bioactive lipids** | **Mean Concentration ± SD** | | | | | | **P Value** |
|---|---|---|---|---|---|---|---|
| | **Recovered (n=30)** | | | **Died (n=20)** | | | |
| AA (µM) | 100.65 | ± | 53.49 | 98.05 | ± | 69.04 | 0.304 |
| LA (µM) | 235.32 | ± | 105.78 | 181.51 | ± | 104.06 | **0.007** |
| DHA (µM) | 40.71 | ± | 23.79 | 38.12 | ± | 31.15 | 0.121 |
| EPA (µM) | 27.64 | ± | 18.88 | 23.87 | ± | 24.97 | 0.064 |
| 18-HEPE (nM) | 10.88 | ± | 13.70 | 9.35 | ± | 14.78 | 0.183 |
| 17-HDHA (nM) | 89.03 | ± | 93.27 | 88.00 | ± | 125.33 | 0.320 |
| 14-HDHA (nM) | 102.49 | ± | 71.29 | 79.05 | ± | 65.09 | 0.218 |
| Maresin 2 (nM) | 0.39 | ± | 0.37 | 0.48 | ± | 0.63 | 0.801 |
| RvD4 (nM) | 4.23 | ± | 3.50 | 3.92 | ± | 4.51 | 0.226 |
| 5,6-EET (nM) | 0.28 | ± | 0.19 | 0.33 | ± | 0.40 | 0.523 |
| 5,6-DHET (nM) | 2.86 | ± | 1.88 | 1.59 | ± | 1.09 | **0.003** |
| 5,6-Ratio | 0.19 | ± | 0.27 | 0.25 | ± | 0.19 | **0.034** |
| 8,9-EET (nM) | 0.65 | ± | 0.64 | 0.89 | ± | 1.13 | >0.999 |
| 8,9-DHET (nM) | 0.45 | ± | 0.28 | 0.50 | ± | 0.36 | 0.658 |
| 8,9-Ratio | 2.62 | ± | 3.63 | 2.64 | ± | 4.72 | 0.911 |
| 11,12-EET (nM) | 170.48 | ± | 132.45 | 146.65 | ± | 104.12 | 0.631 |
| 11,12-DHET (nM) | 0.59 | ± | 0.36 | 0.61 | ± | 0.24 | 0.361 |
| 11,12-Ratio | 399.07 | ± | 465.97 | 308.92 | ± | 291.50 | 0.283 |
| 14,15-EET (nM) | 0.39 | ± | 0.26 | 0.50 | ± | 0.62 | 0.486 |

| | | | | | | | |
|---|---|---|---|---|---|---|---|
| 14,15-DHET (nM) | 0.47 | ± | 0.26 | 0.59 | ± | 0.27 | 0.051 |
| 14,15-Ratio | 0.97 | ± | 0.72 | 0.81 | ± | 0.73 | 0.147 |
| TBXB2 (nM) | 69.61 | ± | 97.96 | 53.91 | ± | 42.15 | 0.864 |
| PGE2 (nM) | 0.55 | ± | 0.37 | 0.57 | ± | 0.48 | 0.593 |
| PGD2 (nM) | 1.04 | ± | 1.29 | 1.25 | ± | 1.41 | 0.967 |
| LTB4 (nM) | 5.27 | ± | 0.57 | 5.36 | ± | 0.78 | 0.837 |
| 5-HETE (nM) | 155.31 | ± | 156.77 | 162.87 | ± | 162.61 | 0.746 |
| 8-HETE (nM) | 3.51 | ± | 2.66 | 3.53 | ± | 4.11 | 0.449 |
| 9-HETE (nM) | 16.49 | ± | 17.37 | 15.93 | ± | 21.25 | 0.340 |
| 11-HETE (nM) | 20.40 | ± | 17.63 | 19.00 | ± | 22.97 | 0.449 |
| 12-HETE (nM) | 171.91 | ± | 148.01 | 148.16 | ± | 117.99 | 0.688 |
| 15-HETE (nM) | 137.76 | ± | 109.79 | 131.33 | ± | 145.66 | 0.320 |
| 16-HETE (nM) | 0.20 | ± | 0.06 | 0.24 | ± | 0.10 | 0.340 |
| 19-HETE (nM) | 176.98 | ± | 190.56 | 207.08 | ± | 210.74 | 0.943 |
| 20-HETE (nM) | 73.50 | ± | 57.86 | 64.25 | ± | 48.96 | 0.631 |
